# Quantification supports amyloid-PET visual assessment of challenging cases: results from the AMYPAD-DPMS study

**DOI:** 10.1101/2024.05.22.24307653

**Authors:** Lyduine E. Collij, Gérard N. Bischof, Daniele Altomare, Ilse Bader, Mark Battle, David Vállez García, Isadora Lopes Alves, Robin Wolz, Rossella Gismondi, Andrew Stephens, Zuzana Walker, Philip Scheltens, Agneta Nordberg, Juan Domingo Gispert, Alexander Drzezga, Andrés Perissinotti, Silvia Morbelli, Christopher Buckley, Valentina Garibotto, Giovanni B. Frisoni, Gill Farrar, Frederik Barkhof, AMYPAD consortium

## Abstract

Several studies have demonstrated the high agreement between routine clinical visual assessment and quantification, suggesting that quantification approaches could support the assessment of less experienced readers and/or in challenging cases. However, all studies to date have implemented a retrospective case collection and challenging cases were generally underrepresented.

**Methods:** In this prospective study, we included all participants (*N*=741) from the AMYPAD Diagnostic and Patient Management Study (DPMS) with available baseline amyloid-PET quantification. Quantification was done with the PET-only AmyPype pipeline, providing global Centiloid (CL) and regional z-scores. Visual assessment was performed by local readers for the entire cohort. From the total cohort, we selected a subsample of 85 cases 1) for which the amyloid status based on the local reader’s visual assessment and CL classification (cut-off=21) was discordant and/or 2) that were assessed with a low confidence (i.e. ≤3 on a 5-point scale) by the local reader. In addition, concordant negative (*N*=8) and positive (*N*=8) scans across tracers were selected. In this sample, (*N*=101 cases: ([^18^F]flutemetamol, *N*=48; [^18^F]florbetaben, *N*=53) the visual assessments and corresponding confidence by 5 certified independent central readers were captured before and after disclosure of the quantification results.

**Results:** For the AMYPAD-DPMS whole cohort, the overall assessment of local readers highly agreed with CL status (κ=0.85, 92.3% agreement). This was consistently observed within disease stages (SCD+: κ=0.82/92.3%; MCI: κ=0.80/89.8%; dementia: κ=0.87/94.6%). Across all central reader assessments in the challenging subsample, global CL and regional z-scores quantification were considered supportive of visual read in 70.3% and 49.3% of assessments, respectively. After disclosure of quantitative results, we observed an improvement in concordance between the 5 readers (κ_*baseline*_=0.65/65.3%; κ_*post-disclosure*_=0.74/73.3%) and a significant increase in reader confidence (*M*_*baseline*_=4.0 vs. *M*_*post-disclosure*_=4.34, *W*=101056, *p*<0.001).

**Conclusion:** In this prospective study enriched for challenging amyloid-PET cases, we demonstrate the value of quantification to support visual assessment. After disclosure, both inter-reader agreement and confidence showed a significant improvement. These results are important considering the arrival of anti-amyloid therapies, which utilized the Centiloid metric for trial inclusion and target-engagement. Moreover, quantification could support determining Aβ status with high certainty, an important factor for treatment initiation.

## Introduction

Recent advances in anti-amyloid immunotherapies and their availability in routine clinical praxis makes it essential to determine amyloid-β (Aβ) status of potentially eligible patients with high certainty^1^. Within this context, quantifying Aβ positron emission tomography (PET) for routine clinical use to support the diagnostic process of neurodegenerative disorders has received great interest over recent years^2^. Several studies have demonstrated the high agreement between routine clinical visual assessment and quantification, suggesting that quantification approaches could support the assessment of less experienced readers and/or in challenging cases^3-7^. However, all studies to date have implemented a retrospective design, which did not allow for direct assessment of the impact of quantification disclosure on visual-based classification of Aβ status and confidence of the assessment. In addition, while most previous studies have speculated on the value of quantification to support particularly challenging cases, these are generally underrepresented and hence require more detailed investigation to support this statement^6-8^.

The three most comprehensive retrospective studies have illustrated the high agreement (86%-96%) between amyloid-PET visual read and several quantification approaches across the three FDA and EMA approved fluorine-18 radiotracers^9-11^. For [^18^F]flutemetamol, an average agreement of 94% between visual read and standard uptake value ratio (SUVr) quantification derived from local non-harmonized quantification pipelines across 5 large clinical studies has been reported^6^. A very similar percentage agreement (i.e., 96.4%) has been reported for [^18^F]florbetaben, where visual read was compared to quantification across 15 different software packages^7^. Finally, in the arguably more ‘real-world’ IDEAS dataset, consisting mostly of [^18^F]florbetapir scans, an 86% concordance between visual read and Centiloid quantification using the robust PET-only Processing (rPOP) pipeline has been demonstrated^12^.

Centiloid quantification has been more widely implemented in recent years, as it brings the tracer-specific SUVr metric to a standardized scale, providing intuitive and across center/tracer generalizable cut points reflecting overall Aβ pathological burden^13^. Neuropathological studies have shown that the earliest detectable amyloid PET signal occurs around 12 CL, while 21-24 CL best discriminates between subjects with none-to-low Aβ plaque burden and those with intermediate-to-high deposition^14, 15^ and 30 CL is indicative of established Aβ burden^16^. Compared to visual positivity, CL cut-offs generally fall in between these values, ranging roughly between 17 CL for highly experienced readers^4^ and 40 CL in a routine clinical setting^17^, though most consistently around 25 CL^4,5,8,12,14^. Considering the robustness of the measure^18^, the Centiloid metric has been widely implemented in AD interventional trials. For example, the Lecanemab (Eisai)^19^ and Donanemab (Eli Lilly)^20^ phase III trials have implemented CL as their primary target engagement outcome and set a negativity threshold (CL<30 and 24.1, respectively) based on this quantification unit. Moreover, in the Donanemab Phase III trial, treatment was stopped if Centiloids were below 11 in a given scan or below 25 in two consecutive ones. Therefore, quantification could also be considered for the discontinuation of anti-amyloid treatment in future clinical routine. It is therefore key to familiarize routine clinical users with quantitative amyloid-PET measures during their diagnostic workup^5^.

We aimed to determine the value of quantification in challenging clinical amyloid-PET cases using a prospective clinical dataset and design. Here, we selected participants from the Amyloid Imaging to Prevent Alzheimer’s disease Diagnostic and Patient Management Study (AMYPAD-DPMS)^21^, who underwent amyloid-PET imaging as part of their diagnostic work-up^22^ and assessed agreement between visual reads performed at each imaging site by ‘local’ readers and quantification performed centrally. For the primary analysis, we selected a subset of challenging cases based on the local readers and assessed the agreement among 5 independent ‘central’ readers before and after disclosure of quantitative results, as well as the confidence in their assessments.

## Methods

### Cohort

Amyloid-PET scans were obtained from the AMYPAD-DPMS randomized controlled trial (*N*=840), which recruited patients across the disease *continuum*, including subjective cognitive decline plus (SCD+), mild cognitive impairment (MCI), or dementia from 8 memory clinics across Europe. A detailed description of the baseline characteristics has been described previously^21^. For the current work, the final disease stage (SCD+, MCI or dementia) and etiological diagnosis (AD, non-AD or not yet achieved [NYA]) during the DPMS observation period were used. All participants gave written informed consent. The trial was registered with EudraCT (2017-002527-21). The study was approved by the CCER (Commission Cantonale d’Ethique de la Recherche) in Geneva Switzerland (#2017-01408).

### Patient selection

All participants with an available baseline amyloid-PET scan that passed quality control (see below) for quantification were included (*N*=741). From this cohort, we selected a subsample of 85 amyloid-PET scans, 1) for which the amyloid status based on the local reader assessment and Centiloid (CL) (cut-off=21, reflecting the lower level of the CL that best discriminates none-to-low and intermediate-to-high Aβ buren^14^) was discordant and/or 2) that were assessed with a low confidence (i.e. ≤3 on a 5-point scale) by the local reader. In addition, VR/CL concordant negative (*N*=8) and positive (*N*=8) scans across tracers and sites to represent real-world negative and positive cases were selected from the total cohort to balance the dataset (*N*=101: ([^18^F]flutemetamol, *N*=48; [^18^F]florbetaben, *N*=53).

### PET acquisition and quantification

Scans were acquired according to the standard protocol for each tracer (i.e. [^18^F]florbetaben/ Neuraceq or [^18^F]flutemetamol/Vizamyl), starting at 90 min. post-injection (p.i.) of 350 MBq (±20%) for florbetaben and 185 MBq (±10%) for flutemetamol and collected in 4 frames of 5 min. each (90 to 110 minutes p.i.). PET images were processed centrally using GEHCs AmyPype PET-only pipeline providing global CL (cortical target mask) and regional z-scores (based on the AAL atlas) for 6 cortical ROIs (frontal, anterior cingulate, posterior cingulate/precuneus, lateral parietal, and lateral temporal cortex). In brief, amyloid-PET images undergo frame to frame alignment and summing and images are spatially normalized to the standard Montreal Neurological Institute (MNI152) space using an adaptive template registration method.^23^ The whole cerebellum was used as reference region. Of note, agreement between AMYPYPE Centiloids and those obtained with the standard Centiloid pipeline has been previously established^24^. Interested parties can request access to the AmyPype software through. For the primary analysis, CL>21 was considered the cut-point for a positive amyloid-PET scan. For illustrative purposes, scans were additionally classified as negative (CL<10), intermediate or so-called ‘gray-zone’ (10>CL<30), and positive (CL>30).

### Visual assessment

Visual assessment was performed according to established reader guidelines; by the local readers for the total cohort and by 5 certified independent central readers for the selected subsample. Images were rated together with an T1-weighted MR scan or CT scan and as either positive (binding in one or more cortical brain region unilaterally, or striatum in case of [^18^F]flutemetamol) or negative (predominantly white matter uptake). In addition, regional\ classifications and reader confidence for both the final and regional visual classification based on a 5-point Likert scale were captured. To assess the effect of quantification disclosure on visual assessment of the 5 central readers of the challenging cases subsample, visual read (VR) and corresponding confidence were captured before and after disclosure of the quantification results. Readers also stated whether CL quantification and/or the regional z-scores supported their assessment or not. Readers were blinded to clinical information.

Importantly, all subsample cohort readers received a short training on the AmyPype processing pipeline, Centiloid quantification anchor points based on the review from Pemberton et al., (2022)^2^, and z-score quantification. The training material can be found in **supplementary materials**.

### Statistical analysis

All analyses were performed in R Studio V4.2.2. Disease stage and etiological diagnostic group differences in quantitative amyloid burden were assessed using ANOVA analysis, corrected for age and sex. Agreement metrics were assessed using Cohen’s or Fleiss Kappa, when applicable. First, agreement between local readers and CL quantification status across the whole cohort and stratified by disease stage was assessed. Next, agreement between local readers and central readers was determined, where a majority VR was established based on the 5 readers (i.e., 3/5 assessments reflected majority Aβ status). Changes in reader confidence after disclosure of quantitative results were assessed using Wilcoxon rank test, as the data was left-skewed.

## RESULTS

The total quantitative cohort consisted of 223 (30.1%) patients with SCD+, 258 (34.8%) with MCI, and 260 (35.1%) with dementia. The mean age was 70.8 years (7.6), 44.8% were female and the average MMSE was 25.5 (4.3). Overall, 49.5% of patients were considered VR-positive based on the local reader assessment (**Table-1**).

**Table 1.** Demographics.

|  | SCD+ |  |  | MCI |  |  | Dementia |  |  | Total Cohort |  |  |
| --- | --- | --- | --- | --- | --- | --- | --- | --- | --- | --- | --- | --- |
|  | AD<br>(N=37) | Non-AD<br>(N=105) | NYA<br>(N=81) | AD<br>(N=119) | Non-AD<br>(N=95) | NYA<br>(N=44) | AD<br>(N=194) | Non-AD<br>(N=49) | NYA<br>(N=17) | AD<br>(N=350) | Non-AD<br>(N=249) | NYA<br>(N=142) |
| **Age, y** | 70.2<br>(5.50) | 68.3<br>(6.21) | 68.7<br>(6.54) | 72.4<br>(6.73) | 70.0<br>(7.92) | 68.1<br>(10.5) | 73.1<br>(7.76) | 71.4<br>(7.96) | 70.5<br>(8.57) | 72.6<br>(7.25) | 69.6<br>(7.32) | 68.7<br>(8.15) |
| **Sex, F(%)** | 15<br>(40.5%) | 42<br>(40.0%) | 39<br>(48.1%) | 54<br>(45.4%) | 36<br>(37.9%) | 21<br>(47.7%) | 102<br>(52.6%) | 16<br>(32.7%) | 7<br>(41.2%) | 171<br>(48.9%) | 94<br>(37.8%) | 67<br>(47.2%) |
| **MMSE** | 28.4<br>(1.71) | 28.6<br>(1.43) | 28.9<br>(1.48) | 25.9<br>(2.72) | 26.2<br>(3.42) | 26.5<br>(2.88) | 21.4<br>(4.53) | 24.2<br>(3.74) | 23.6<br>(4.13) | 23.7<br>(4.59) | 26.8<br>(3.28) | 27.6<br>(2.97) |
| **VR+ (%)** | 37<br>(100%) | 13<br>(12.4%) | 9<br>(11.1%) | 106<br>(89.1%) | 13<br>(13.7%) | 5<br>(11.4%) | 178<br>(91.8%) | 3<br>(6.1%) | 3<br>(17.6%) | 321<br>(91.7%) | 29<br>(11.6%) | 17<br>(12.0%) |
| **Centiloid** | 75.0<br>(34.1) | 9.85<br>(23.9) | 11.3<br>(28.3) | 75.9<br>(37.5) | 12.0<br>(30.8) | 10.3<br>(30.1) | 85.6<br>(38.7) | 8.47<br>(33.8) | 12.1<br>(34.3) | 81.2<br>(38.1) | 10.4<br>(28.7) | 11.1<br>(29.4) |

*SCD+:* subjective cognitive decline plus; *MCI:* mild cognitive impairment; *AD:* Alzheimer's disease; *NYA:* not yet achieved; *MMSE:* mini-mental state examination; *VR+:* visual read positivity

The ‘challenging’ subsample included mostly MCI patients (52 [51.5%], followed by SCD+ (29 [28.7%]), and finally dementia (20 [19.8%]). The mean age of this subpopulation was 72.5 years (7.6), 44.6% were female and the average MMSE was 26.2 (4.3). Overall, 57.4% of patients were considered VR-positive based on the local reader assessment (**Table-2**).

**Table 2.** Demographics of challenging subsample cohort.

|  | **SCD+**<br>(N=29) | **MCI**<br>(N=52) | **Dementia**<br>(N=20) | **Whole Cohort**<br>(N=101) |
| --- | --- | --- | --- | --- |
| **Age** | 67.93 (5.7) | 73.87 (7.2) | 75.40 (5.9) | 72.47 (7.2) |
| **Sex (F)** | 10 (34.5%) | 26 (50.0%) | 9 (45.0%) | 45 (44.6%) |
| **MMSE** | 28.6 (1.6) | 26.2 (3.2) | 22.9 (4.5) | 26.2 (3.7) |
| **Centiloid** | 32.8 (29.8) | 41.9 (31.8) | 27.1 (25.5) | 36.3 (30.4) |
| **VR+ (%)** | 14 (48.3%) | 33 (63.5%) | 11 (55.0%) | 58 (57.4%) |

*SCD+*: subjective cognitive decline plus; *MCI*: mild cognitive impairment; *MMSE*: mini-mental state examination; *VR+*: visual read positivity

### Quantitative amyloid burden across diagnostic groups

Global amyloid burden expressed in CL showed a stepwise increase with disease stage (SCD+<MCI<dementia: *F*=60.5, *p*<0.001, **Table-1, Figure-1A**) and was higher in AD than in non-AD or “not yet achieved” etiological diagnostic groups (*F*=411.9, *p*<0.001, **Table-1, Figure-1B**). However, amyloid burden did not differ across the different clinical disease stages within etiological groups (**Supplementary Figure-1**). Regional z-scores were highest in the AD group (*p*_all_<0.01) but did not differ between the non-AD and “not yet achieved” group.

**Figure 1.**
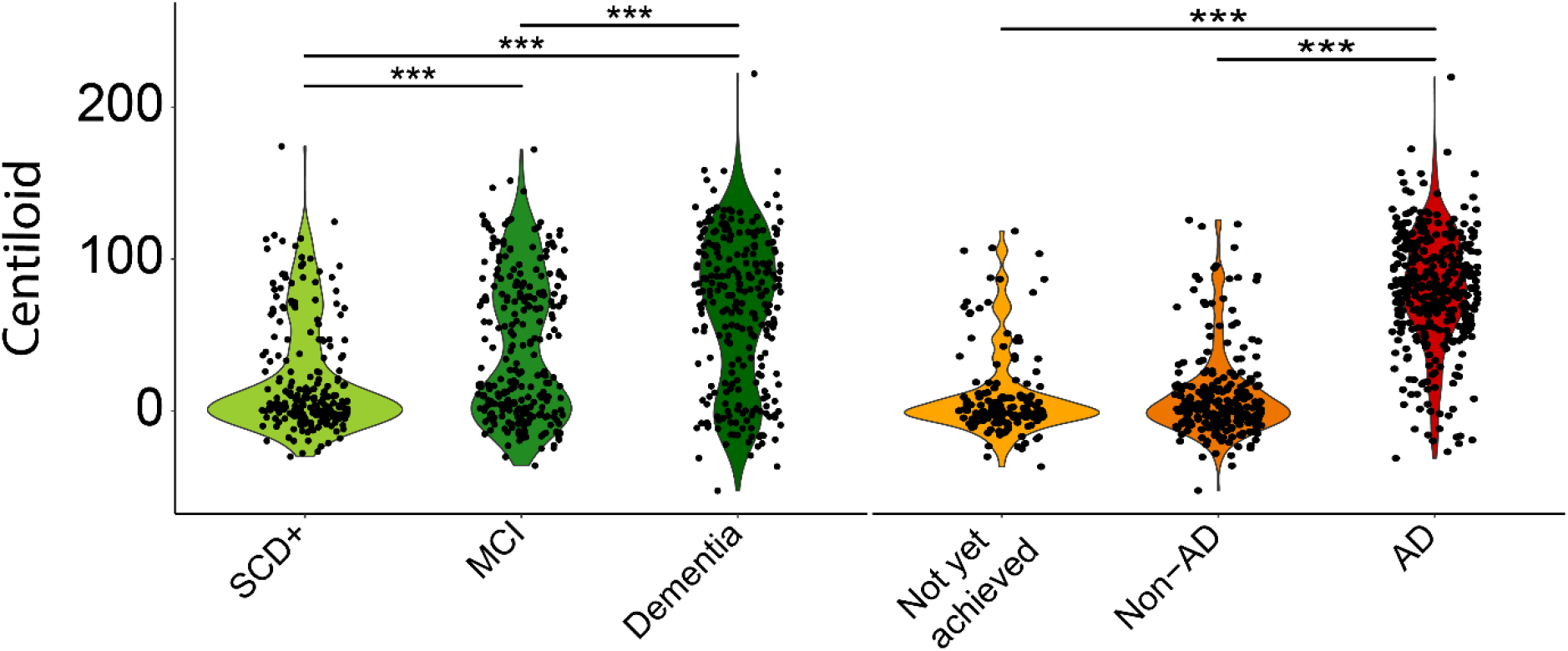
Centiloid quantification across disease stages and etiological diagnosis. Violin plot shows the distribution of Centiloid burden across disease stages (left panel) and etiological diagnosis (right panel). Significant differences between groups are illustrated, after correction for age and sex.

### Agreement between local readers and Centiloid quantification

For the whole quantitative cohort (*N*=741), the overall assessment of local readers highly agreed with CL status based on the predefined cutoff of 21 (κ=0.85, 92.3% concordance). This high agreement was consistently observed within disease stage, ranging from κ=0.87/94.6% for dementia cases, κ=0.82/92.3% for SCD+, and κ=0.80/89.8% for MCI (**Figure-2**).

**Figure 2.**
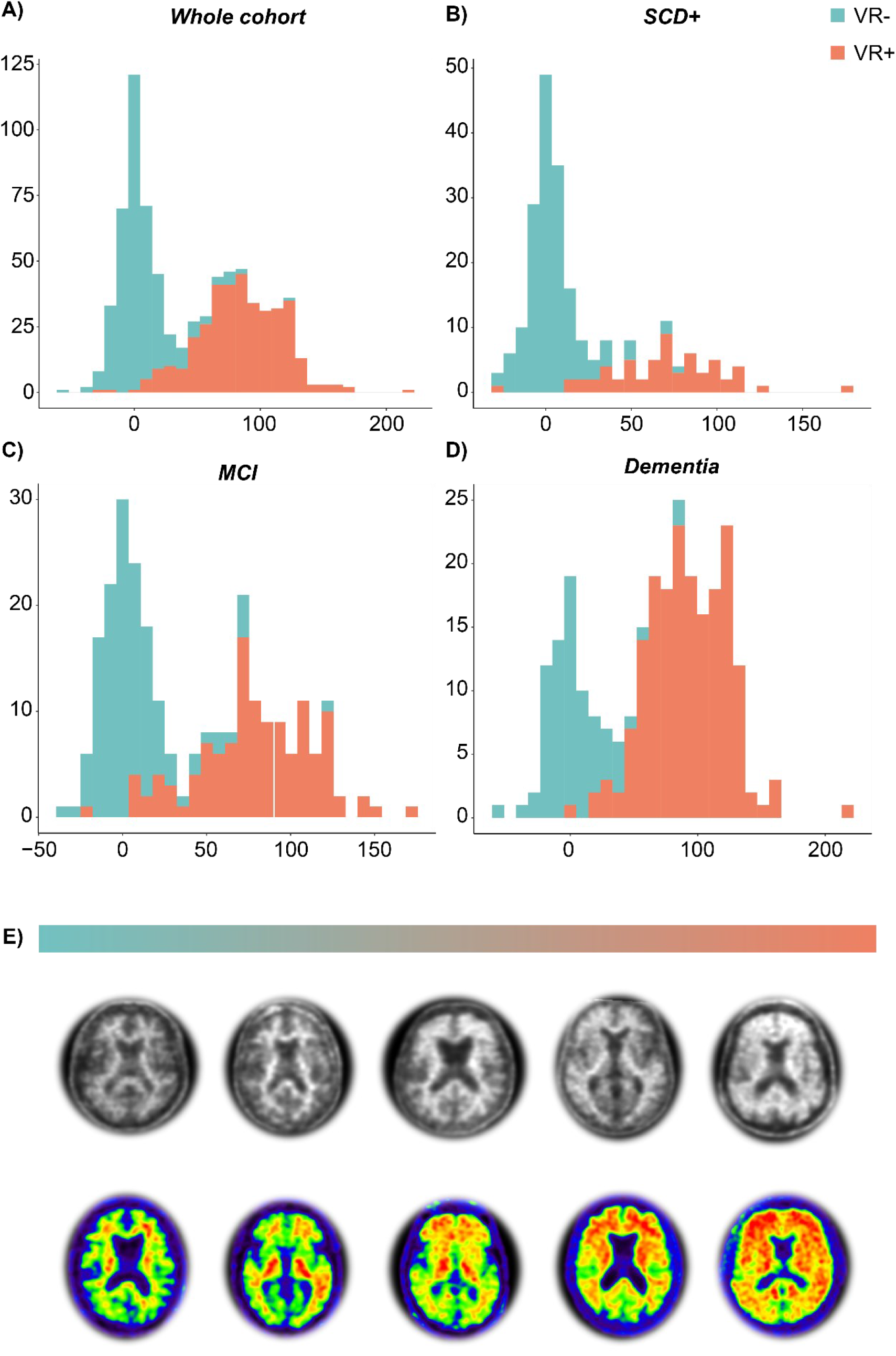
Agreement between local visual read and Centiloid quantification in the whole DPMS cohort. Histograms illustrate the distribution of Centiloid across **A)** the whole quantitative DPMS cohort, **B)** the SCD+ patient population, **C)** the MCI patient population, and **D)** the dementia patient population. Bars are color coded for visual read status by the local assessor. **E)** Illustrative amyloid-PET from negative (left) to global positive (right). Top row represents [^18^F]florbetaben and bottom row represents [^18^F]flutemetamol scans.

### Local vs Central readers

For the subsample enriched with challenging cases (*N*=101), the agreement between local and central readers was as per study design low (κ=0.21). Importantly, with majority central read as the reference standard, local readers were more inclined to classify an amyloid-PET scan as negative, resulting in 29 (28.7%) false-negative cases and 12 (11.9%) false-positive cases.

### Quantification supports visual read of challenging cases

Across all 5 certified independent central reader assessments (*N*=505), global CL and regional z-score quantification was considered supportive of VR in 70.3% and 49.3% of assessments, respectively. **Figure-3** illustrates changes in number of positive visual assessments (0-5 as per number of readers) pre- and post-disclosure of quantitative results. After disclosure of the quantitative results, we observed an improvement in concordance between the 5 readers (κ_*baseline*_=0.65/65.3%; κ_*post-disclosure*_=0.74/73.3%), which can be appreciated in the **Figure-3** post-disclosure column, where relatively more cases were consistently VR- or VR+ for all readers. In line, we also observed a slight improvement in agreement between the consensus read and amyloid status based on CL (κ_*baseline*_=0.53; κ_*post-disclosure*_=0.60). Finally, a significant increase in reader confidence (*M*_*baseline*_=4.0 vs. *M*_*post-disclosure*_=4.34, *W*=101056, *p*<0.001) after disclosure of quantitative results was observed.

**Figure 3.**
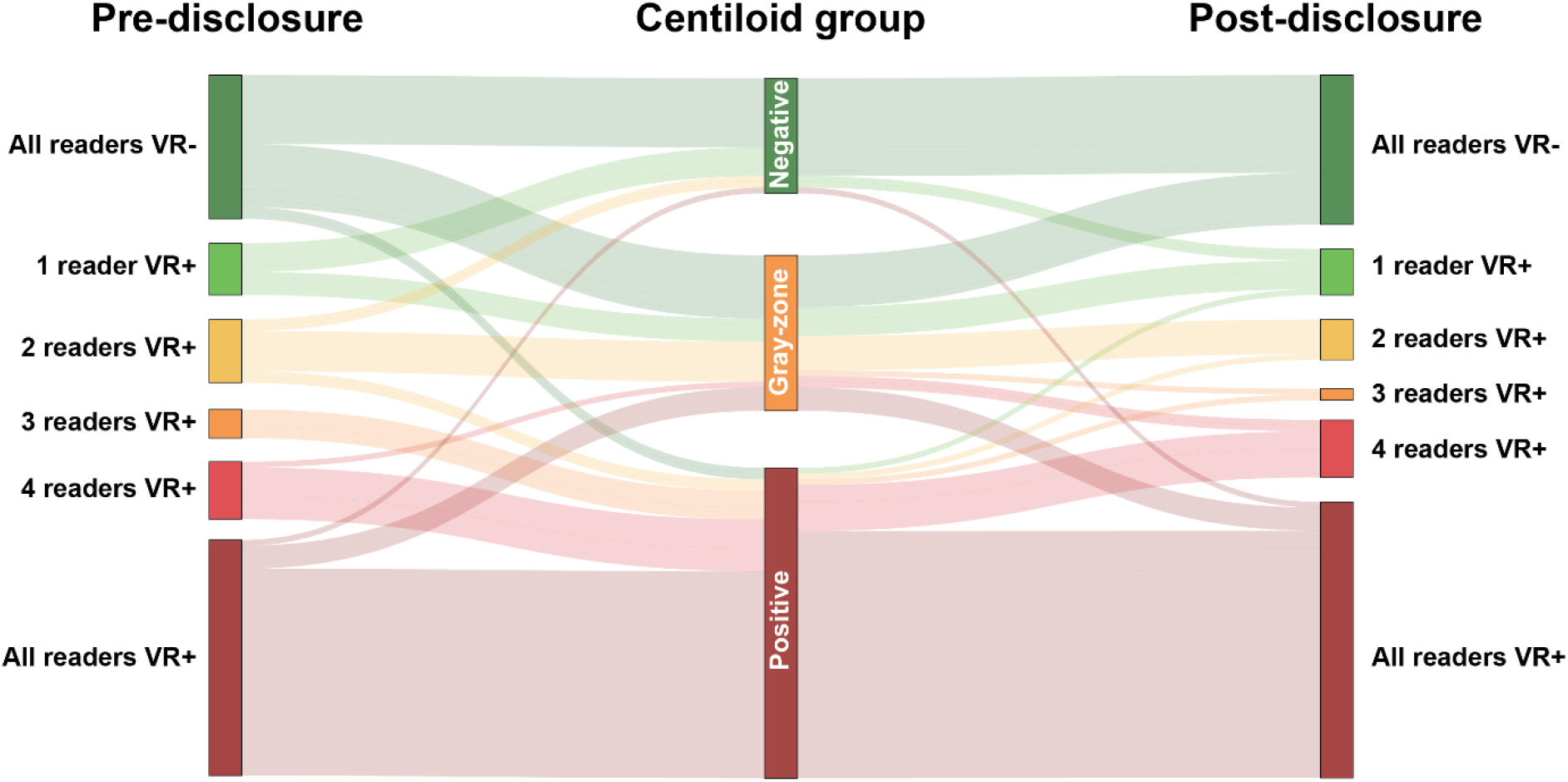
Change in visual read status after disclosure of quantitative results. Sankey plot illustrates changes in the number of positive visual reads pre- and post-disclosure of quantitative results. Change is illustrated by Centiloid group, *i*.*e*., Negative (CL<10), Gray-zone (10>CL<30), and Positive (CL>30). Note, these CL group classifications were not shared with the reader but were created post-hoc based on the literature for visualization purposes.

Nonetheless, some cases did not reach consensus between readers or showed clear discrepancy between visual assessment and Centiloid quantification. Examples are illustrated and further commented on in **Figure-4**.

**Figure 4.**
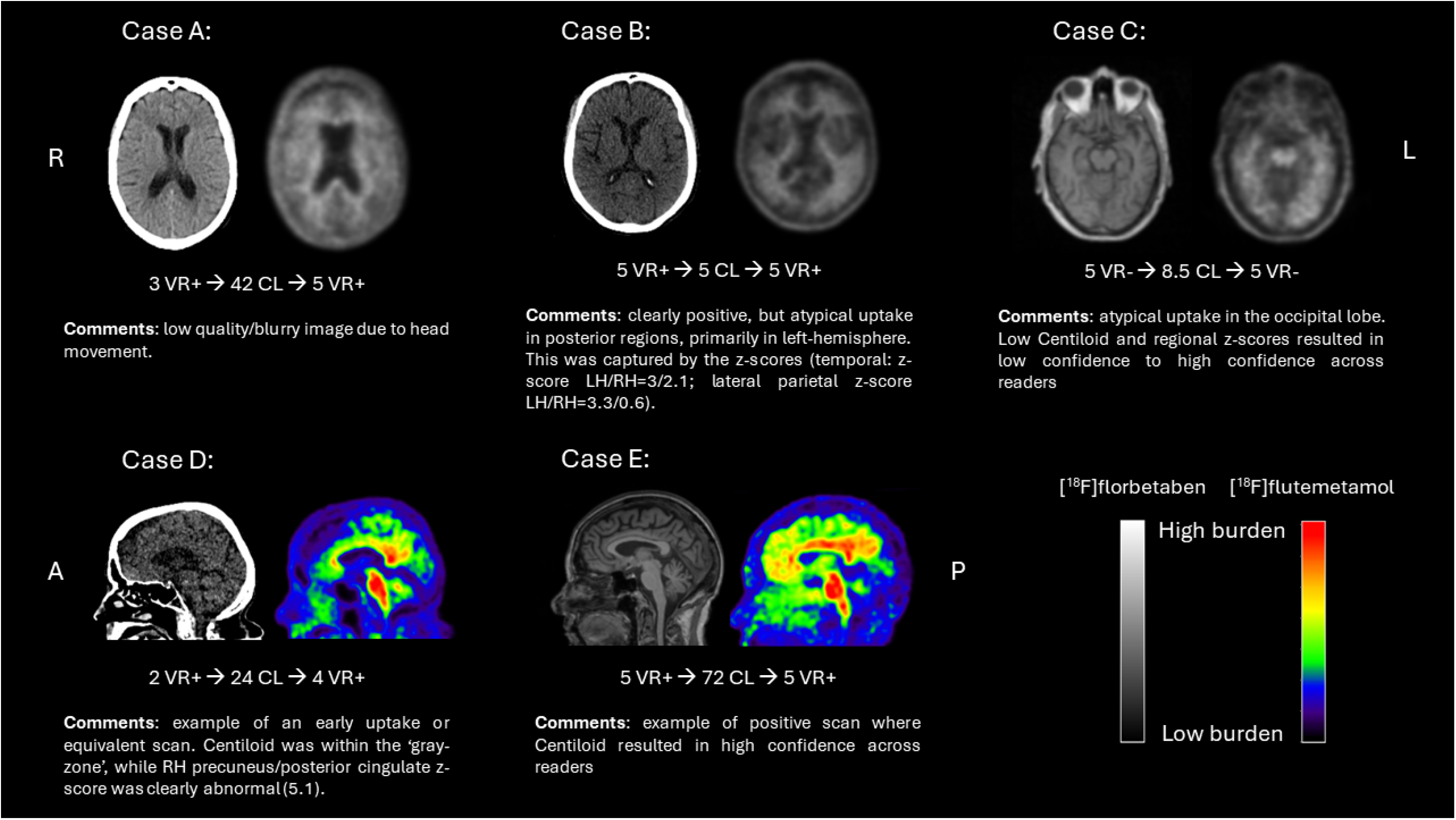
Examples of challenging cases. R/RH: right hemisphere; L/LH: left hemisphere; A: anterior orientation; P: posterior orientation; CL: Centiloid; VR: visual read

### Possible additional value of regional z-scores

As stated above, for 249/505 of the central assessments the regional z-scores were considered helpful in addition to the global CL quantification. This was more apparent for 3 out of 5 raters, who stated the regional z-scores added to their read in 74, 67, and 62 (out of 101) cases, compared to the 24 and 22 cases for the other 2 raters. For 48 cases across central readers more detailed comments were provided, which suggested that the main benefit of regional z-scores was in case of a borderline scan (28/48, 58.3%) and particularly for the frontal and PC/PCC regions, followed by quality of the image and/or atrophy (8/48, 16.7%). In 8 instances (16.7%), the regional z-score caused confusion rather than further support of the initial visual assessment.

## DISCUSSION

In the prospective DPMS study, we observed excellent agreement between local visual reads and CL quantification across the clinical *continuum*. In a subgroup enriched for challenging cases we demonstrate that an improvement in reader agreement and confidence can be achieved by using quantification results. While overall agreement between local readers and quantification is high and consistent across cognitive stages, approximately 11% of scans in the AMYPAD-DPMS cohort representative of a typical memory clinical population was considered challenging due to a variety of causes, such as suboptimal scan quality, atrophy, or emerging Aβ pathology/borderline scan. In the case of the latter, regional z-scores might be of additional support to the global CL metric.

Our results suggest that quantification can support readers in determining Aβ status with high certainty, which is crucial considering the arrival of anti-amyloid therapies and their associated costs and potential side-effects. Similar to previous studies^6,12, 25^, approximately 8% of AMYPAD-DPMS cases showed discordance between local readers and CL quantification. Though the CL cut-off utilized in the current work was at the lower end of the range that best discriminates none-to-low and intermediate-to-high Aβ buren^14^, some considerable discrepancies were observed (**Figure-2**). These examples possibly reflect misdiagnosis and could explain some of the previously reported discrepancies between visual read status and final diagnosis for this cohort^22^. It is important to note, that even though the potential value of quantification has been previously demonstrated^3-7^, most studies probably underestimated its true impact in a real-world scenario, due to its limited value when visual assessment yields a clear positive or negative outcome. This study did evaluate amyloid-PET quantification performance in a wide range of amyloid load from negative to positive (local reads) and in a subset of challenging cases where quantification seems more beneficial as visual analysis alone can be insufficient or less accurate. Nonetheless, quantification should always be done in conjunction with visual assessment, to avoid misclassifications due to potential quantification errors. For example, one case had a very low CL value, but was consistently assessed as visual read positive by all readers (**Figure-3&4 case B**).

In addition to high certainty in Aβ status, the extent of burden as expressed in CL units also has clinical relevance considering the inclusion and discontinuation of treatment criteria implemented in the two successful anti-amyloid trials. More specifically, the Lecanemab Phase III trial defined amyloid-positivity as a CL>30, while Donanemab utilized a CL>37 cut-off. The real-world IDEAS study demonstrated that around two thirds of the discordant cases were assessed as visually positive but classified as amyloid negative based on CL^12^. In a future era of anti-amyloid therapies, the adjunctive use of quantification could avoid such false-positive patients being unnecessarily medicated for treatment regimens which potentially could last 1-2 years without any therapeutic value but with the risk of side effects. While quantification is already added to the label of both radiotracers used in this study by the EMA, current FDA guidelines for amyloid-PET do not mention the added value that quantitation could bring to reaching high confidence and accurate determination of Aβ-status based on visual reads. In addition, as the clearance rate for Donanemab was so high, the study also implemented a treatment discontinuation criterion, namely when the amyloid-PET quantification was CL<11 or CL<25 in two consecutive scans. To what extent these specific cut-offs will be implemented in the user-criteria for lecanemab and donanemab remains to be determined, as the current appropriate use recommendations (*e*.*g*., for lecanemab^26^) only elude to a ‘positive amyloid-PET or CSF result indicative of AD’. Nonetheless, some initial results suggest a steady clearance rate of Aβ, independent of baseline amyloid burden^27^. As such, future work should investigate whether the extent of baseline Aβ burden is predictive of necessary treatment duration to achieve full Aβ clearance. In such a setting, quantification will not only inform on Aβ status, but also optimize individual treatment plans.

A limitation of the study is that we did not repeat visual assessments by the local readers after disclosure of quantitative results. Though our central readers also had different levels of experience, this might have been ever more dispersed across 11 site local readers. Also, we had limited data to investigate whether subjects in the CL grey-zone would convert to a Aβ-positive status at follow-up.

## CONCLUSION

In this prospective study and subsample enriched for challenging amyloid-PET cases, we demonstrate the value of quantification to support visual assessment. After disclosure, inter-reader agreement and confidence showed a significant improvement. These results are important considering the arrival of anti-amyloid therapies, which utilize the Centiloid metric for trial inclusion and target-engagement. Moreover, quantification could support determining Aβ status with high certainty, an important feature for treatment initiation.

## Data Availability

Data is available upon request through the ADDI platform

https://amypad.eu/

## ACKNOWLEDGMENTS

The project leading to this paper has also received funding from the Innovative Medicines Initiative 2 Joint Undertaking under grant agreement No 115952. This Joint Undertaking receives the support from the European Union’s Horizon 2020 research and innovation programme and EFPIA. This communication reflects the views of the authors and neither IMI nor the European Union and EFPIA are liable for any use that may be made of the information contained herein.

## REFERENCES

1. Perneczky R, Jessen F, Grimmer T, et al. Anti-amyloid antibody therapies in Alzheimer’s disease. Brain 2023;146:842–849.

2. Pemberton HG, Collij LE, Heeman F, et al. Quantification of amyloid PET for future clinical use: a state-of-the-art review. Eur J Nucl Med Mol Imaging 2022;49:3508–3528.

3. Collij LE, Konijnenberg E, Reimand J, et al. Assessing Amyloid Pathology in Cognitively Normal Subjects Using (18)F-Flutemetamol PET: Comparing Visual Reads and Quantitative Methods. J Nucl Med 2019;60:541–547.

4. Collij LE, Salvado G, Shekari M, et al. Visual assessment of [(18)F]flutemetamol PET images can detect early amyloid pathology and grade its extent. Eur J Nucl Med Mol Imaging 2021;48:2169–2182.

5. Collij LE, Salvado G, de Wilde A, et al. Quantification of [(18) F]florbetaben amyloid-PET imaging in a mixed memory clinic population: The ABIDE project. Alzheimers Dement 2023;19:2397–2407.

6. Bucci M, Savitcheva I, Farrar G, et al. A multisite analysis of the concordance between visual image interpretation and quantitative analysis of [(18)F]flutemetamol amyloid PET images. Eur J Nucl Med Mol Imaging 2021;48:2183–2199.

7. Jovalekic A, Roe-Vellve N, Koglin N, et al. Validation of quantitative assessment of florbetaben PET scans as an adjunct to the visual assessment across 15 software methods. Eur J Nucl Med Mol Imaging 2023.

8. Zeltzer E, Mundada NS, La Joie R, et al. Quantitative analysis of 6,150 real-world amyloid Positron Emission Tomography (PET) scans from the Imaging Dementia–Evidence for Amyloid Scanning (IDEAS) study. Alzheimer’s & Dementia 2022;18:e066217.

9. Barthel H, Gertz HJ, Dresel S, et al. Cerebral amyloid-beta PET with florbetaben (18F) in patients with Alzheimer’s disease and healthy controls: a multicentre phase 2 diagnostic study. Lancet Neurol 2011;10:424–435.

10. Curtis C, Gamez JE, Singh U, et al. Phase 3 trial of flutemetamol labeled with radioactive fluorine 18 imaging and neuritic plaque density. JAMA Neurol 2015;72:287–294.

11. Clark CM, Schneider JA, Bedell BJ, et al. Use of florbetapir-PET for imaging beta-amyloid pathology. JAMA 2011;305:275–283.

12. Iaccarino L, La Joie R, Koeppe R, et al. rPOP: Robust PET-only processing of community acquired heterogeneous amyloid-PET data. Neuroimage 2022;246:118775.

13. Klunk WE, Koeppe RA, Price JC, et al. The Centiloid Project: standardizing quantitative amyloid plaque estimation by PET. Alzheimers Dement 2015;11:1–15 e11-14.

14. La Joie R, Ayakta N, Seeley WW, et al. Multisite study of the relationships between antemortem [(11)C]PIB-PET Centiloid values and postmortem measures of Alzheimer’s disease neuropathology. Alzheimers Dement 2019;15:205–216.

15. Amadoru S, Dore V, McLean CA, et al. Comparison of amyloid PET measured in Centiloid units with neuropathological findings in Alzheimer’s disease. Alzheimers Res Ther 2020;12:22.

16. Salvadó G, Molinuevo JL, Brugulat-Serrat A, et al. Centiloid cut-off values for optimal agreement between PET and CSF core AD biomarkers. Alzheimer’s Research & Therapy 2019;11:27.

17. Hanseeuw BJ, Malotaux V, Dricot L, et al. Defining a Centiloid scale threshold predicting long-term progression to dementia in patients attending the memory clinic: an [(18)F] flutemetamol amyloid PET study. Eur J Nucl Med Mol Imaging 2021;48:302–310.

18. Shekari M, Salvadó G, Battle MR, et al. Evaluating robustness of the Centiloid scale against variations in amyloid PET image resolution. Alzheimer’s & Dementia 2021;17:e055726.

19. van Dyck CH, Swanson CJ, Aisen P, et al. Lecanemab in Early Alzheimer’s Disease. New England Journal of Medicine 2022;388:9–21.

20. Sims JR, Zimmer JA, Evans CD, et al. Donanemab in Early Symptomatic Alzheimer Disease: The TRAILBLAZER-ALZ 2 Randomized Clinical Trial. JAMA 2023;330:512–527.

21. Altomare D, Collij L, Caprioglio C, et al. Description of a European memory clinic cohort undergoing amyloid-PET: The AMYPAD Diagnostic and Patient Management Study. Alzheimers Dement 2022.

22. Altomare D, Barkhof F, Caprioglio C, et al. Clinical Effect of Early vs Late Amyloid Positron Emission Tomography in Memory Clinic Patients: The AMYPAD-DPMS Randomized Clinical Trial. JAMA Neurol 2023;80:548–557.

23. Lundqvist R, Lilja J, Thomas BA, et al. Implementation and validation of an adaptive template registration method for 18F-flutemetamol imaging data. J Nucl Med 2013;54:1472–1478.

24. Collij LE, Farrar G, Vallez Garcia D, et al. The amyloid imaging for the prevention of Alzheimer’s disease consortium: A European collaboration with global impact. Front Neurol 2022;13:1063598.

25. Jovalekic A, Roé-Vellvé N, Koglin N, et al. Validation of quantitative assessment of florbetaben PET scans as an adjunct to the visual assessment across 15 software methods. European Journal of Nuclear Medicine and Molecular Imaging 2023;50:3276–3289.

26. Cummings J, Apostolova L, Rabinovici G, et al. Lecanemab: appropriate use recommendations. The journal of prevention of Alzheimer’s disease 2023;10:362–377.

27. Lilly scientific information - Clinical Trials on Alzheimer’s Disease (CTAD) 2023. https://medical.lilly.com/us/science/conferences/neuroscience/ctad.

