## supplement for "Quantification supports amyloid-PET visual assessment of challenging cases: results from the AMYPAD-DPMS study"

**Supplementary Materials**

**Methods – Training materials readers**

The following figures were presented to the central readers as training material for the current study:

**1. Centiloid anchor points**

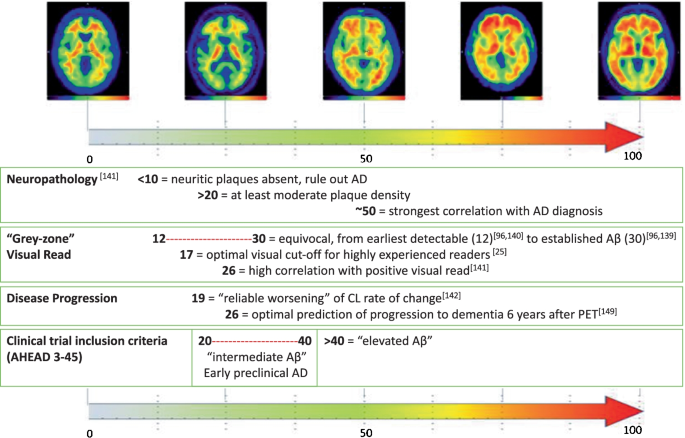

Adapted from Pemberton et al., 2022 *European Journal of Nuclear Medicine and Molecular Imaging* 49(10):3508-3528. doi: 10.1007/s00259-022-05784-y

**2. AmyPype processing pipeline**

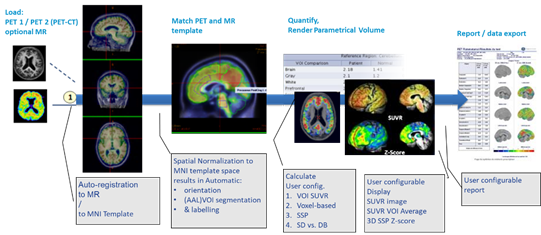

Courtesy of GE Healthcare

**3. Example quantification outputs**

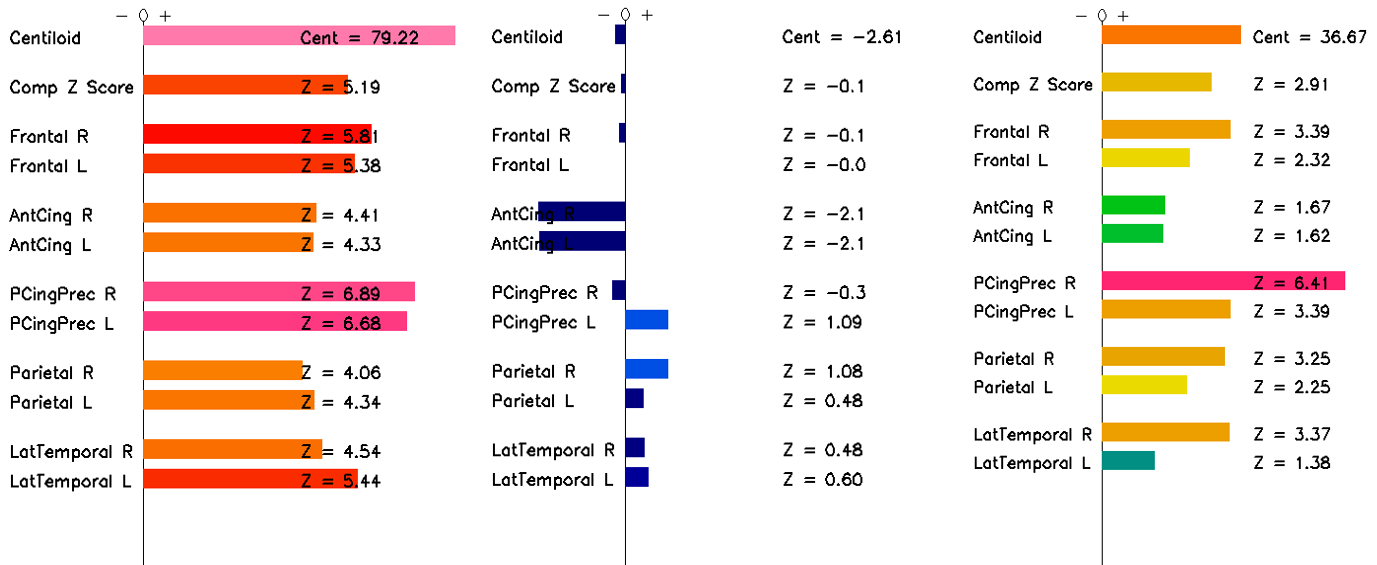

Courtesy of GE Healthcare

**Results**

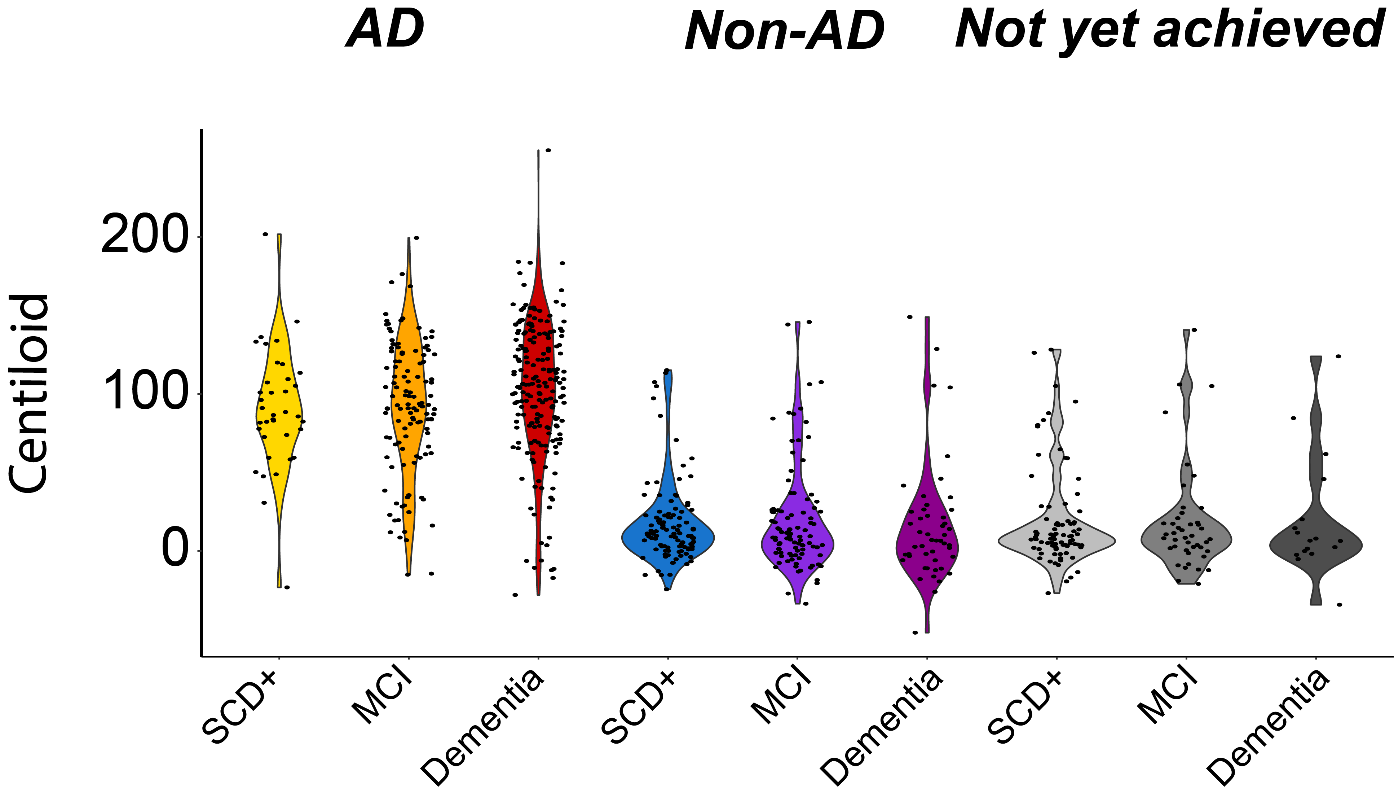

**Supplementary Figure 1. Centiloid quantification across syndromic and etiological groups**

Violin plot illustrates the distribution of quantitative amyloid-PET burden, as expressed in Centiloid units, across the whole population stratified by syndromic diagnostic group and per etiological group.

*SCD+: subjective cognitive decline plus; MCI: mild cognitive impairment; AD: Alzheimer’s disease*
